# HIV and Hepatitis B virus co-infection in Mozambique: policy review and health professionals’ knowledge and practices

**DOI:** 10.1101/2024.03.23.24304747

**Authors:** Vanda dos Muchangos, Lúcia Chambal, Charlotta Nilsson, Esperança Sevene

## Abstract

**Background:** Human Immunodeficiency Virus (HIV) and Hepatitis B Virus (HBV) co-infection is a public health problem affecting 2.7 million worldwide. In Mozambique, the prevalence of this co-infection is 9.1%, calling for specific policies on prevention, diagnosis and adequate management in health facilities caring for HIV patients. This study aimed to review the existing policies and to assess the knowledge and practices of health professionals about HIV/HBV co-infection.

**Methods:** A document and literature review to describe the existing policies and guidelines on HIV/HBV co-infection in Mozambique was performed. Key informants were contacted to clarify or add information. Health Professionals who care for HIV-positive patients in four health centers in Maputo City, the capital of Mozambique, responded to a questionnaire on knowledge and practices about this co-infection. Qualitative analysis was done to identify main themes using content analysis. Descriptive statistics of socio-demographic, knowledge and practice variables was presented using the SPSS Program version 20 and bivariate analysis was applied to describe the association between variables.

**Results:** Twenty-one policy documents were found, and five key informants were interviewed. Fifty-two participants answered the questionnaire. Only one policy document explicitly referred to HIV/HBV co-infection treatment. Most Health Professionals (96%) were aware of HIV/HBV co-infection. Although the only existing policy is on the treatment, few (33%) referenced antiretroviral formulations containing Tenofovir and Lamivudine. Only 29% of Health Professionals reported screening HIV patients for HBV and 21% practiced HIV/HBV co-infection counselling. No statistically significant differences were found when relating the socio-demographic variables with knowledge and practices.

**Conclusion:** Policy documents relating to prevention, diagnosis and clinical management of HIV/HBV co-infection were rare or absent. Health Professionals had little knowledge about HIV/HBV co-infection. Defining adequate policies and training of Health Professionals may help increase awareness, increase counselling of patients for disease prevention, diagnosis and proper management of HIV/HBV co-infected patients.

## Introduction

According to UNAIDS (2023), around 39 million people globally were living with human immunodeficiency virus (HIV) in 2022 of which 37.5 million were adults and 1.5 million children. Fifty-three % of all people living with HIV (PLWH) were women and girls. Eastern and southern Africa remains the region most heavily affected by HIV, with 20.6 million PLWH—54% of all PLWH. In Mozambique, the HIV prevalence is 13.2% (1).

With the introduction of specific policies, including policies for access to care and antiretroviral treatment, the average life expectancy of PLWH has been increasing. With increasing longevity, there is an increase in morbidity and mortality associated with other pathologies in patients with HIV, including hepatitis B virus infection (HBV) (2)(3). In 2019, 296 million people were living with chronic hepatitis B worldwide (4), with a higher prevalence of chronic HBV infection and higher rates of HBV-associated hepatocellular carcinoma (HCC) in sub-Saharan Africa (5)(6). Studies have shown a HBV infection prevalence in Mozambique at 10.6% (7)(8) and 12.2% (8). HBV can lead to chronic infection with progression to liver cirrhosis, HCC, liver failure, and death, and the patients with chronic infection become carriers and transmitters of HBV infection (9).

HBV and HIV share the same transmission routes, which include (a) exposure of mucous membranes or non-intact skin to contaminated blood and body fluids (saliva, semen, vaginal fluids), (b) contact with exudates from lesions and contaminated surfaces, (c) interpersonal contact through sharing contaminated objects (needles, syringes, toothbrushes, razors), and (d) mother-to-child transmission. HBV represents a growing cause of mortality and morbidity in HIV co-infected patients, including patients on ART (10)(11).

HIV infection negatively affects the natural course of HBV-induced hepatitis (12). Co-infected patients are less likely to resolve the acute HBV infection spontaneously, have a higher risk of progression to liver complications with fibrosis (13) and tend to develop cirrhosis and HCC in less time compared to HBV mono-infected patients (14). Other consequences of HIV/HBV co-infection include reduced response to treatment, cross-drug resistance to HIV and HBV, and increased risk of drug hepatotoxicity (15)(16).

According to the 2021 WHO Global progress report on HIV, viral hepatitis and sexually transmitted infections, in 2015 2.7 million people were co-infected with HIV and hepatitis B virus, and of these, 1.9 million (69% of all cases) lived in sub-Saharan African countries (4)(17). The prevalence of HIV/HBV co-infection in Mozambique in 2015 was 9.1% (18). A study of HCC patients at Maputo Central Hospital showed a prevalence of HBV and HIV of 56.1% and 18.6%, respectively (19), highlighting the importance of integrating HBV testing in providing care and attention to HIV-positive patients. This approach would benefit from the infrastructure of HIV care, and it would improve co-infected patients’ health conditions and treatment and reduce morbidity and mortality resulting from co-infection (18).

WHO recommends HBsAg testing to all groups considered at high risk for HBV, including HIV-positive patients, relatives and sexual contacts of patients with HBV, men who have sex with men, sex workers, and prisoners. This recommendation is critical to ensure that non-immune risk groups are vaccinated. It also recommends that HBsAg-positive individuals be referred to evaluate other markers of hepatitis B and staging liver disease for adequate care. The WHO also recommends that if treatment is needed as per HIV or HBV parameters, the ART regimen should contain at least two drugs active against HIV and HBV. The ART regimen would preferably include Tenofovir (TDF) and Emtricitabine (FTC) or Lamivudine (3TC) (20).

To implement the WHO recommendations, Health Professionals (HP) must be guided by clear policies or guidelines (19)(21)(22), have adequate training and have knowledge about the management of this co-infection to adjust their practices. Lack of knowledge about HBV and inappropriate practices were described in low- and middle-income countries (23)(24) and high-income countries (25). In Mozambique, there is little information on policies for prevention and management of co-infected patients. This study aimed to evaluate the knowledge and practices of HPs about HIV/HBV co-infection.

## Methods

### Study design and settings

The study took place from January to May 2019 using mixed methods with qualitative and quantitative approaches. First, a document and literature review to describe the existing policies and guidelines on HIV/HBV co-infection was performed. Second, key informants were in-depth interviewed to complement, clarify or add information. After the policy review, a cross-sectional study was carried out. HPs who care for HIV-positive patients in four health centers (HC) in Maputo City, the capital of Mozambique, responded to a semi-structured questionnaire on knowledge and practices about HIV and HBV co-infection.

The municipality of Maputo is the largest city in Mozambique and the country’s administrative, political, economic and cultural capital, with a population of 1 101170 according to the National population Census of 2017 (26). It is divided into seven Municipal Districts: Ka’Mpfumo, Nlhamankulu, Ka’Maxakeni, Ka’Mubukwana, Ka’Mavhota, Ka’Tembe and Ka’Nyaka, where the health centers are located. The most populous district is Ka’Mavhota, with just over a quarter of the city’s total population, followed by the Nlhamankulu district, with just under a quarter. The prevalence of HIV in Maputo City is 16% (1). Maputo City has 31 HCs that serve PLWH distributed over the city’s seven districts. Five HCs were selected based on the number of HIV patients on antiretroviral treatment. The Mavalane HC is located in Ka’Mavhota district, Alto-Maé HC in Ka’Mpfumo, Polana Caniço and 1° de Maio HCs in Ka’Maxakeni. In these HCs, the services for PLWH are organized into adult outpatient clinics, maternal and child health clinics that include care for pregnant women and HIV-positive children, and counselling services for HIV testing and treatment.

### Study population and tools

The key informants were heads of relevant programs and institutions involved in the health policy development process in the country, the National Blood Service, Blood Bank, National Directorate of Pharmacy, National Program for the Control of STIs-HIV/AIDS and Hepatitis, and the Immunization Program. For the interview, a script with systematized questions was used to identify documents that refer to policies and regulations regarding the prevention, diagnosis and treatment of HBV-infection in HIV-positive patients. The script contained a section with identification and socio-demographic data, questions regarding the type of policy document, the year of publication, key information regarding prevention, diagnosis and treatment, issuing authority and the document’s source.

The HPs were medical doctors (general practitioners), medical technicians (general medicine technicians), nurses (with medium and higher training in nursing) and counselors (with basic training in counselling for testing and treatment of HIV patients) in service for more than six months in HIV-positive patient care. They worked in outpatient clinics for chronic illness, maternal and child health care, prenatal consultation, counselling and testing. They were selected because were involved in the WHO cascade to cure viral hepatitis B and C framework used in this study. Care is given in three levels: first, the counsellors provide counselling for HIV prevention and testing; second, the nurses carry out testing for diagnosis, counselling and follow-up of the patient’s treatment; third, the medical technicians and medical doctors make the diagnosis, prescribe the medicines, follow-up the patient and reinforce prevention measures. Participants were selected for convenience according to the distribution and operation of services for HIV-positive patients in the four health units.

Each participant answered a semi-structured questionnaire divided into five-sections, including sociodemographic characteristics, HBV and HIV/HBV co-infection knowledge and practices regarding handling patients with HIV/HBV co-infection, and participant vaccination status (S1). The questions were closed with one open option at the end to include any additional information from the participant. The questions were revised to adapt to the participants’ comprehension based on a pre-test.

### Data collection

For the desk review, relevant official documents from the Ministry of Health (MoH) were reviewed. The documents were obtained through the government portal and MoH website, using keywords: Hepatitis B, HIV/hepatitis B co-infection, policy, norm, Mozambique, and some documents were provided in physical or electronic format by key informants. The literature review was done electronically using PubMed, Google Academic and Hinari databases. For the search, the words Mozambique with hepatitis B, HIV/HBV co-infection, policies, norms and HIV/HBV co-infection with policies and norms were cross-referenced. Each document was reviewed in a systematized manner, and documents in Portuguese or English were included from the period of January 1977 to February 2019. Only articles available in the full text were reviewed. After the literature search, all abstracts were extracted and screened. The relevant papers and the documents obtained via key informants and desk review were included in this study (PRISMA diagram). After explaining the study procedure and respecting confidentiality and privacy, all participants voluntarily gave written informed consent. Key informants were approached by the principal investigator at their working sectors to conduct a 30 minutes interview focused on the existing policies for the prevention, diagnosis and treatment of HIV and HBV co-infected patients. It included information regarding the documents found in the desk review, the capture of other relevant documents and clarification of possible points of divergence from the information reviewed.

The researcher interviewed HPs for approximately 45-60 minutes at the participants’ most convenient time without workload. The questionnaires were in Portuguese, the local national language.

### Data management and analysis

The policy documents were organized according to type, year and document source, key information regarding policies on HBV prevention, diagnosis and treatment in HIV co-infected patients. Policy documents from the MoH and articles were reviewed to describe policies. Each document was reviewed in a systematized manner to identify the type of document, the name and year of publication, and key policy information for the prevention, diagnosis and management of hepatitis in HIV/HBV co-infected patients.

A content analysis of the policy documents regarding the prevention, diagnosis and management of HIV/HBV co-infection was performed. The content of the documents was transcribed to Excel, where they were tabulated according to the predefined themes for describing policies on prevention, diagnosis and treatment of HIV/HBV co-infection patients. The analysis theme was based on the WHO "Monitoring and Evaluation of Viral Hepatitis B and C" Framework (14) for eliminating viral hepatitis as a public health threat by 2030, which describes the ten core indicators for monitoring and evaluation of viral hepatitis B and C. It includes indicators related to the cascade for a cure, such as prevention, testing, diagnosis, treatment, and cure (outputs and outcomes).

New themes that emerged were added to the document table. For Knowledge and practices about HIV and HBV co-infection among health professionals data were collected in a questionnaire paper form and then entered into a database on Open Data Kit (ODK) (27) in a tablet. After cleaning, the data was imported into the Scientific Package for Health Sciences (SPSS) Program in version 20 for analysis.

To ensure data quality, a daily review of each questionnaire was carried out to assess their consistency and completeness. All out-of-bounds values were reviewed and corrected. This review was carried out during the data collection process and after entering the data. A descriptive analysis of the socio-demographic, knowledge and practice variables was presented. A bivariate analysis of the association between the knowledge and practice variables and the socio-demographic variables (gender, professional experience, profession) was carried out using the Pearson Chi-square test (X²) using a 5% statistical significance level.

### Ethic consideration

The Institutional Bioethics Committee for Health of the Faculty of Medicine/Maputo Central Hospital (CIBS FM & HCM/068/2018) and the Scientific Council of the Faculty of Medicine at Eduardo Mondlane University approved the study protocol. The study researchers were responsible for ensuring that the protocol, IC and data collection instruments were reviewed and approved by the Ethics Review Committees.

#### Informed Consent

The Informed Consent (IC), written in Portuguese, was given by the study investigator to each eligible participant. The participant was given time to read and understand the content of the IC and to consult someone they trusted about the information received before deciding to consent. The researcher clarified any doubts that arose regarding objectives and confidentiality. Inclusion in the study was voluntary and only took place after signing the Informed Consent form. Participants were able to withdraw from the study at any time. All participants gave written informed consent.

## Results

### Policy review

Forty documents were identified, 36 resulting from the document and literature review and four provided by key informants. Twenty-one policy documents were included in the study, 17 national and four international. Nineteen documents were excluded because they were not related to HIV and HBV co-infection policies and were not related to Mozambique. We found ordinances, decrees, strategic plans, treatment guides, and resolutions. Figure 1 presents the prism diagram adapted from Moher D, 2009.

**Figure 1.**
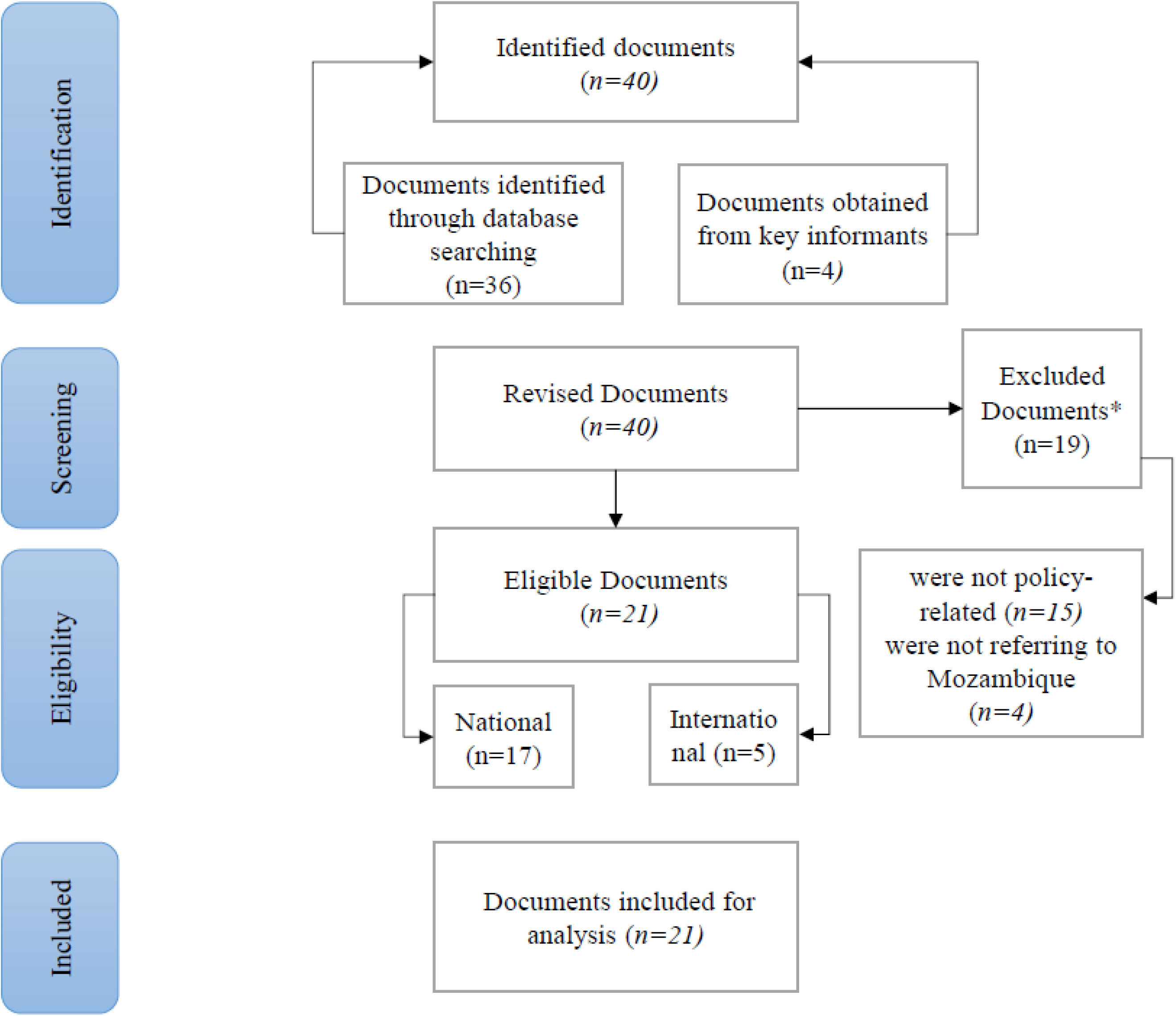
Document Review Diagram. Source: Adapted from Prism Diagram. Moher D, 2009

The documents were established from 1977 and 2019 by the Ministry of Health, Government of Mozambique and WHO, with key information regarding prevention, diagnosis and treatment of HBV in HIV patients (Table S1).

A key informant from each of the five sectors of origin of the documents was interviewed, namely, from the Blood Bank, National Directorate of Pharmacy, National Program for the Control of STIs-HIV/AIDS and Hepatitis, Immunization Program, and National Blood Service. The key informants were heads of the program or their representatives with an average length of service of 14 years (5 to 27 years). The median age of the five respondents was 44 years, of which three were women.

No specific policy related to HIV/HBV co-infection was found, and recommendations concerning these diseases were in documents related to other policies. The majority of policies were linked to hepatitis B prevention. The biosafety policies were present in different documents, and it has “Safe Injections” through observation of all procedures associated with the preparation of injections, the use of disposable syringes and needles and the proper disposal of material used in the injection. “Blood safety” for the prevention of blood-borne infections for a donation, which includes testing for four infections (HIV, HBV/HCV, Syphilis) and selection of low-risk donors based on a questionnaire, was also legislated (table 1). A key informant stated that:

> *Our principle for blood safety is that for all blood-borne diseases, not just Hepatitis B, the first filter is a selection through a questionnaire with exclusion questions for those who have risk factors or risk behaviours for the transmission of Hepatitis B in this case…(Key informant I)*

**Table 1.** Existing policies regarding prevention, diagnosis and treatment of Hepatitis B.

| **Policy** | **Year** | **Description** |
| --- | --- | --- |
| **Prevention** |  |  |
| Blood Safety Policy | 1977 | Prevention of blood borne infections for donation (HBV/HIV testing) |
| HBV Vaccination Policy | 2001 | Prevention of HBV infection in children under 1 year of age |
| Prophylactic treatment policy for all healthcare professionals exposed to HIV | 2001 | Prevention of HIV Infection to the Healthcare Professional |
| Injection Safety Policy | 2002 | Infection prevention (HIV/HBV) in clients and Health Professionals with the introduction of disposable syringes, one needle and one syringe for each patient, observance of procedures in the preparation of injections and disposal of used injectable material |
| HBV infection prevention policy for all exposed Health professionals and victims of sexual assault | 2009 | HBV vaccination to exposed Health professionals and victims of sexual assault after exposure, in the 1st and 6th month |
| **Diagnosis** |  |  |
| HBV testing policy for all exposed Health Professionals and rape victims | 2014 | HBV testing at Health Professionals after exposure and rape victims |
| **Treatment** |  |  |
| Treatment policy with lines of therapy containing TDF, Lamivudine to HIV/HBV co-infected patients. | 2016 | Consider as co-infected patients those with HbsAg + for more than 6 months<br>Recommendation for treatment of co-infected patients |

HIV- human immunodeficiency virus, HBV-hepatitis B virus, HbsAg-hepatitis B surface antigen, TDF-tenofovir.

In 2006, the MoH introduced the HBV vaccine, which is administered to children under one year of age as part of a pentavalent vaccine (DPT-Hepb-Hib) at month 2, month 3 and month 4 of age. In 2009, MoH recommended vaccination against HBV for adults, adolescents and pregnant women who were victims of rape. In 2014, the guidelines were updated to include children after a rape and health professionals after exposure. The HBV vaccine is administered immediately after the event, in the 1st and 6th months, in a total of 3 doses. These policies do not include routine vaccination of risk groups such as health professionals, HIV positive patients, drug users and neonates (Table 1).

> *Well, this is a pentavalent vaccine, in addition to Hepatitis B, there are others, DTP, Hepatitis B, ….so it is a conjugate vaccine, so it is not a single Hepatitis B vaccine… we are thinking of giving the vaccine to the Newborn, but it’s still a plan… (Key informant II)*

Regarding the diagnosis of HBV infection, we did not find specific regulations. The 2014 Guide for Antiretroviral and Opportunistic Infections Treatment recommends testing for HBV in victims of rape and health professionals after exposure. The Guide does not refer to the diagnosis of HBV in HIV positive patients or other risk groups (men who have sex with men, sex workers, intravenous drug users) (table 1). On the other hand, key informants referred to tests performed for screening of blood donors.

> *Few patients arrive at the Central Hospital If we had a routine test for Hepatitis in our public network, we would have more patients in follow-up, but we don’t, so those patients who are in a disease phase arrive at the Central Hospital can no longer be helped, and those who are referred from our blood donation services (Key informant III)*

Regarding the treatment of HBV infection, the guide for Antiretroviral and Opportunistic Infections Treatment (2016) recommends that the co-infected patient be treated with a therapeutic line containing HBV active drugs (TDF+3TC+EFV). It also recommends strictly monitoring liver and kidney function avoiding alcohol and traditional medicines consumption due to the risk of hepatotoxicity. The National Drug Formulary of September 2007 indicates combinations with Lamivudine for the Treatment of HIV and chronic HBV infection. Key informants mentioned that only HIV/HBV co-infected patients benefit from treatment:

> *These patients end up benefiting from HIV treatment, so we still don’t have a follow-up norm; at the moment, what we are doing here as Ministry of Health, we are preparing our national guide for the screening, diagnosis, treatment of patients with Hepatitis. We have a technical group working on it. (Key informant III)*

### Health professionals’ knowledge and practice

For the assessment of knowledge and practices, 52 HPs were interviewed. The mean age was 35 years old (Min 23-Max 58), and 40 (77%) were female. All participants were trained in in management of HIV/AIDS, in contrast, only 7 (14%) had received training related to HBV. Seventy-one % of participants reported having received at least one dose of the HBV vaccine. However, only 11 (21%) received all three doses, and only 9 (17%) knew how many doses were needed to provide immunity against the infection. “Table 2” presents the sociodemographic characteristics, vaccination status and distribution of the participants by health facility and training in HBV.

**Table 2.**
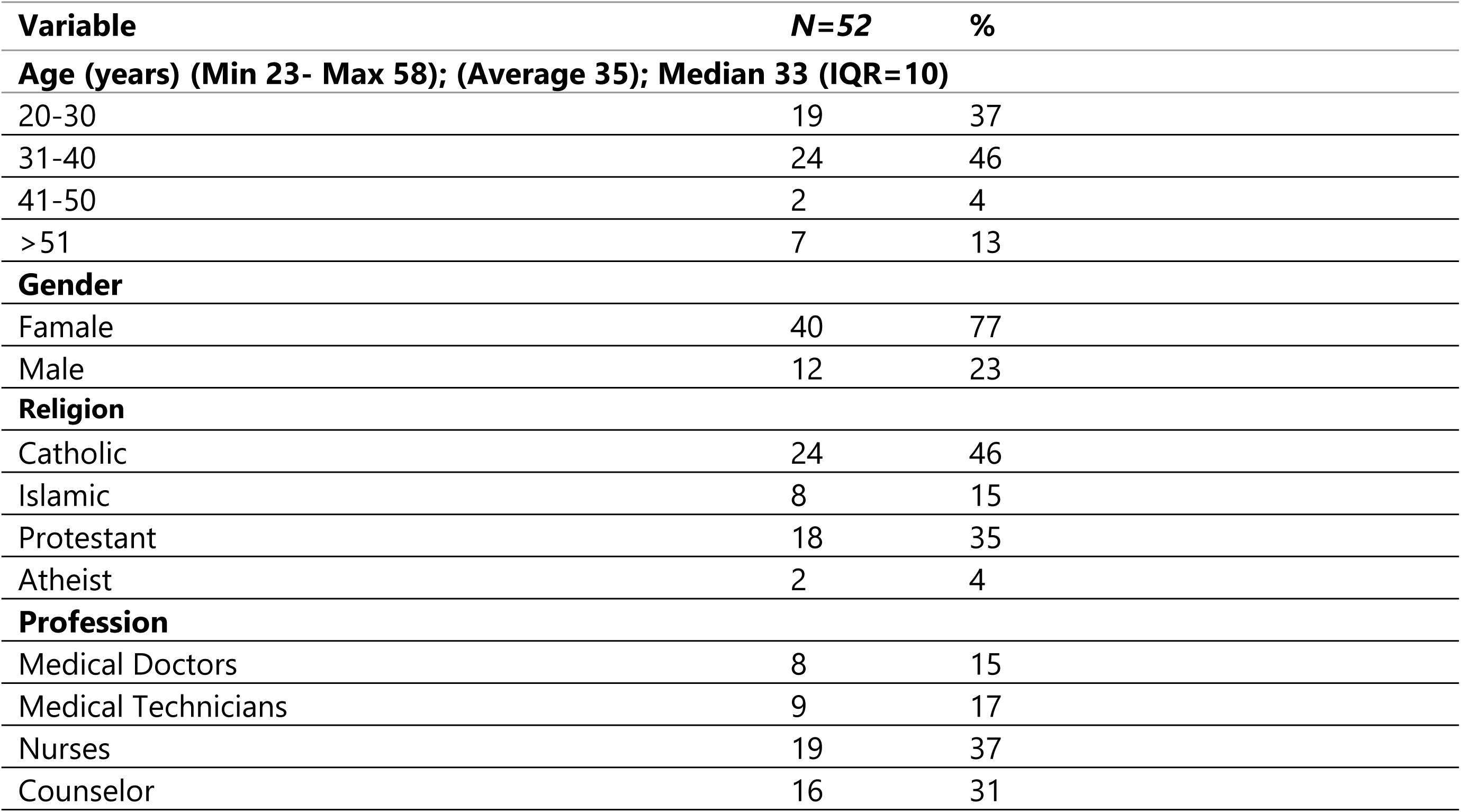
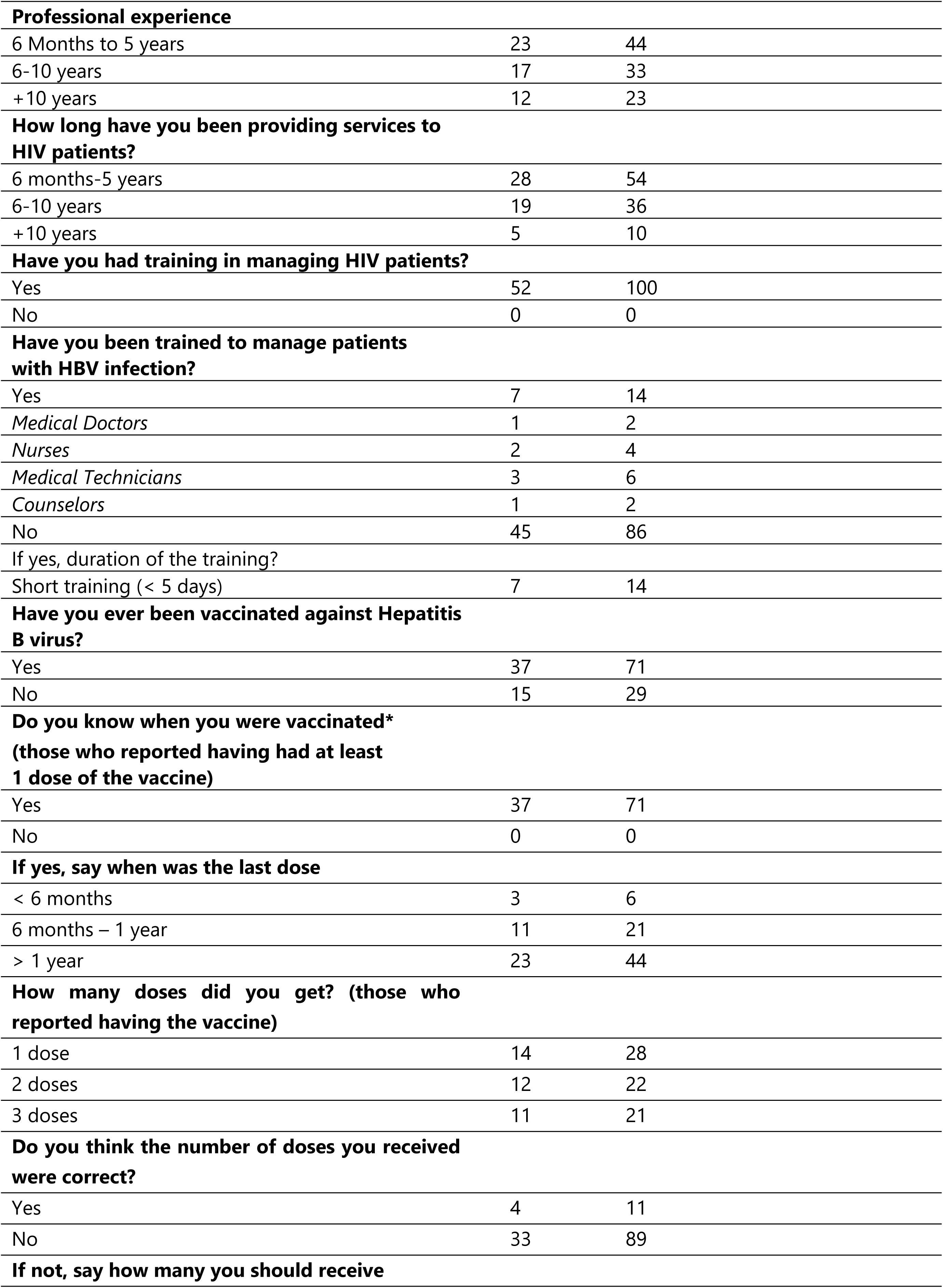
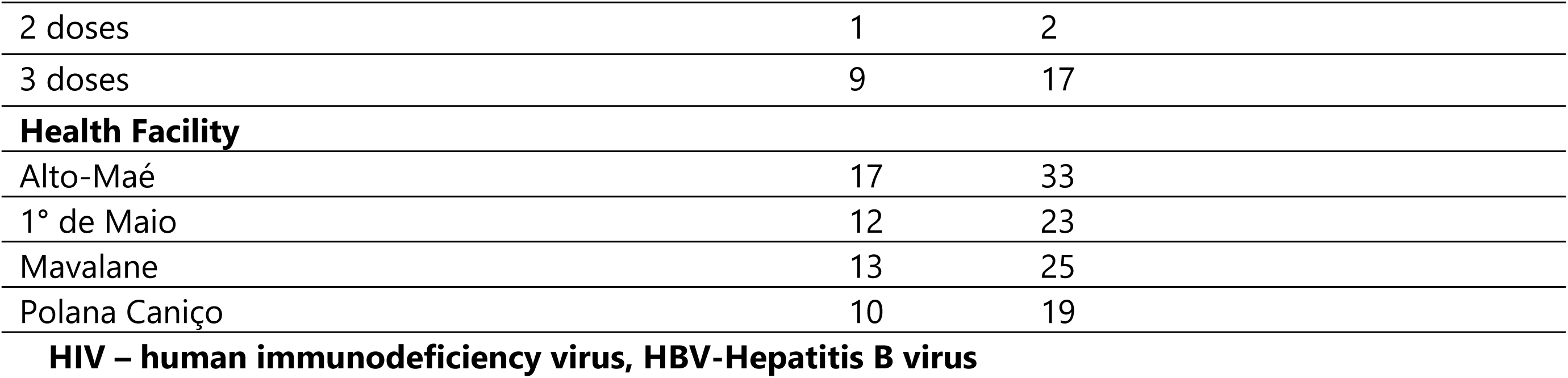
Sociodemographic characteristics, vaccination status and distribution of participants by health facility and training in HBV.

| **Variable** | **N=52** | **%** |
| --- | --- | --- |
| **Age (years) (Min 23- Max 58); (Average 35); Median 33 (IQR=10)** |  |  |
| 20-30 | 19 | 37 |
| 31-40 | 24 | 46 |
| 41-50 | 2 | 4 |
| >51 | 7 | 13 |
| **Gender** |  |  |
| Famale | 40 | 77 |
| Male | 12 | 23 |
| **Religion** |  |  |
| Catholic | 24 | 46 |
| Islamic | 8 | 15 |
| Protestant | 18 | 35 |
| Atheist | 2 | 4 |
| **Profession** |  |  |
| Medical Doctors | 8 | 15 |
| Medical Technicians | 9 | 17 |
| Nurses | 19 | 37 |
| Counselor | 16 | 31 |
| **Professional experience** |  |  |
| 6 Months to 5 years | 23 | 44 |
| 6-10 years | 17 | 33 |
| +10 years | 12 | 23 |
| **How long have you been providing services to HIV patients?** |  |  |
| 6 months-5 years | 28 | 54 |
| 6-10 years | 19 | 36 |
| +10 years | 5 | 10 |
| **Have you had training in managing HIV patients?** |  |  |
| Yes | 52 | 100 |
| No | 0 | 0 |
| **Have you been trained to manage patients with HBV infection?** |  |  |
| Yes | 7 | 14 |
| *Medical Doctors* | 1 | 2 |
| *Nurses* | 2 | 4 |
| *Medical Technicians* | 3 | 6 |
| *Counselors* | 1 | 2 |
| No | 45 | 86 |
| If yes, duration of the training? |  |  |
| Short training (< 5 days) | 7 | 14 |
| **Have you ever been vaccinated against Hepatitis B virus?** |  |  |
| Yes | 37 | 71 |
| No | 15 | 29 |
| **Do you know when you were vaccinated\* (those who reported having had at least 1 dose of the vaccine)** |  |  |
| Yes | 37 | 71 |
| No | 0 | 0 |
| **If yes, say when was the last dose** |  |  |
| < 6 months | 3 | 6 |
| 6 months – 1 year | 11 | 21 |
| > 1 year | 23 | 44 |
| **How many doses did you get? (those who reported having the vaccine)** |  |  |
| 1 dose | 14 | 28 |
| 2 doses | 12 | 22 |
| 3 doses | 11 | 21 |
| **Do you think the number of doses you received were correct?** |  |  |
| Yes | 4 | 11 |
| No | 33 | 89 |
| **If not, say how many you should receive** |  |  |
| 2 doses | 1 | 2 |
| 3 doses | 9 | 17 |
| **Health Facility** |  |  |
| Alto-Maé | 17 | 33 |
| 1° de Maio | 12 | 23 |
| Mavalane | 13 | 25 |
| Polana Caniço | 10 | 19 |

**HIV – human immunodeficiency virus, HBV-Hepatitis B virus**

### Knowledge about HBV infection and HIV/HBV co-infection

In the evaluation of knowledge, 98% of the participants already had information about HBV infection. Regarding the mode of transmission, 38 (73%) reported shared sharp objects, 25 (48%) participants mentioned sexual contact, 15 (29 %) blood transfusion and 10 (19%) mother-to-child transmission. Regarding the prevention methods, 18 (35%) reported using condoms, and only 7 (14%) of the participants mentioned vaccination to prevent HBV. None of the medical technicians or counselors described the vaccine as a means of prevention.

Regarding diagnosis, only 8 (15%) described that the HBsAg marker is used to screen for HBV.

Regarding treatment, 12 (33%) have information that hepatitis B is treatable; of these, 5 (14%) medical doctors, 5 (14%) medical technician and 2 (5%) nurses described that Tenofovir (TDF) is the drug indicated for the treatment of HBV infection. No medical technician described TDF as the drug indicated for the treatment of HBV infection. In the group of doctors, 2 (25%) said they did not know about the drug indicated for the treatment of HBV infection. In the group of nurses, only one reported that TDF is the drug indicated for the treatment of HBV infection. Counselors were excluded from this analysis, as they do not receive training on treatment modalities.

Regarding knowledge about HBV and HIV co-infection, 50 (96%) of the participants reported knowing that HBV and HIV can exist in the same patient. Only 12 (33%) knew about HBV complications in the co-infected patient (Cirrhosis 11, hepatocellular carcinoma 6, Chronic Hepatitis 2 and Hepatotoxicity to ARVs 1) (table 3). Only 12 (33%) of the participants knew about HBV treatment in patients with HIV and the therapeutic lines indicated for its treatment (it includes two active drugs, Tenofovir and Lamivudine). Counsellors were once again excluded. “Table 3” presents the Knowledge about Hepatitis B and HIV/HBV co-infection.

**Table 3.** Knowledge about Hepatitis B infection and HIV/HBV co-infection.

| Variable | Yes, n (%) | No, n (%) | Total, n (%) |
| --- | --- | --- | --- |
| Have you ever heard of hepatitis B? | 51 (98) | 1 (2) | 52 (100) |
| **Modes of transmission** |  |  |  |
| Sexual contact | 25 (48) | 27 (52) | 52 (100) |
| Sharing needles and syringes | 22 (42) | 30 (58) | 52 (100) |
| Blade Sharing | 16 (31) | 36 (69) | 52 (100) |
| Transfusion | 15 (29) | 37 (71) | 52 (100) |
| Mother to child | 10 (19) | 42 (81) | 52 (100) |
| Saliva | 3 (6) | 49 (94) | 52 (100) |
| Toothbrush sharing | 0 | 52 (100) | 52 (100) |
| Tattoos, piercings, scarifications | 0 | 52 (100) | 52 (100) |
| Breast milk | 0 | 52 (100) | 52 (100) |
| **Modes of prevention** |  |  |  |
| Condom use | 18 (35) | 34 (65) | 52 (100) |
| *Medical Doctors* | 6 (11,5) | NA | NA |
| *Medical Technicians* | 4 (7,7) | NA | NA |
| *Nurses* | 6 (11,5) | NA | NA |
| *Counselors* | 2 (3,8) | NA | NA |
| Safe transfusion | 8 (15) | 44 (85) | 52 (100) |
| Vaccination | 7 (14) | 45 (86) | 52 (100) |
| *Medical Doctors* | 2 (3,9) | NA | NA |
| *Medical Technicians* | 0 | NA | NA |
| *Nurses* | 5 (9,6) | NA | NA |
| *Counselors* | 0 | NA | NA |
| Proper disposal of needles | 7 (14) | 45 (86) | 52 (100) |
| Proper disposal of syringes | 6 (12) | 46 (88) | 52 (100) |
| Use of mask | 2 (4) | 50 (96) | 52 (100) |
| Use of gloves | 1 (2) | 51 (98) | 52 (100) |
| Proper disposal of gloves | 1 (2) | 51 (98) | 52 (100) |
| **Diagnosis is done by** |  |  |  |
| HBsAg | 8 (15) | 44 (85) | 52 (100) |
| *Medical Doctors* | 3 (6) | NA | NA |
| *Medical Technicians* | 3 (6) | NA | NA |
| *Nurses* | 1 (2) | NA | NA |
| *Counselors* | 1 (2) | NA | NA |
| Jaundice | 1 (2) | 51 (98) | 52 (100) |
| Hepatomegaly | 1 (2) | 51 (98) | 52 (100) |
| Blood test | 1 (2) | 51 (98) | 52 (100) |
| **Do you know if HIV and HBV can exist in the same patient?** | 50 (96) | 2 (4) | 52 (100) |
| **Have you ever heard of the treatment of HBV in HIV patients?** | 12 (33) | 24 (67) | 36 (100) |
| *Medical Doctors* | 5 (14) | NA | NA |
| *Medical Technicians* | 5 (14) | NA | NA |
| *Nurses* | 2 (5) | NA | NA |
| **If yes, which medications are indicated?** |  |  |  |
| TDF+3TC+EFV | 12 (33) | 0 | 12 (33) |
| Emtricitabina+Lamivudina | 0 | 12 (33) | 12 (33) |
| **Do you know complications of HBV in patients with HIV?** | 12 (33) | 40 (77) | 52 (100) |

| If yes, what are the complications? |  |  |  |
| --- | --- | --- | --- |
| Cirrhosis | 11 (30) | 1 (2) | 12 (23) |
| Hepatocellular Carcinoma | 6 (16) | 6 (12) | 12 (23) |
| Chronic Hepatitis | 2 (4) | 10 (19) | 12 (23) |
| Hepatotoxicity to ARV | 1 (2) | 11 (21) | 12 (23) |

**HIV**-human immunodeficiency virus, **HBV**-hepatitis B virus, **HBsAg**-hepatitis B surface antigen, **TDF**-tenofovir, **3TC**-lamivudine, **EFV**-efavirenz, **ARV**-antiretrovirals, **NA**-not applicable

### Knowledge about HIV and HBV co-infection according to sociodemographic characteristics

We did not find an association between knowing whether HIV and HBV can exist in the same patient and gender (p=0.412), profession (p=0.197) or professional experience (p=0.269). However, we found an association between knowledge about complications and profession (p=0.001), where medical doctors had more knowledge. It was observed that there was an association between knowing that there is HBV treatment in patients with HIV and the professional category (p=0.009), where medical doctors and medical technicians had more knowledge. This association was not observed with gender (p=0.433) and professional experience (p=0.345) “Table 4”. Counsellors were excluded from this analysis as they do not treat patients.

**Table 4.** Knowledge about co-infection and sociodemographic characteristics.

|  | Do you know that HIV and HBV can exist in the same patient? |  |  |  | Have you ever heard of HBV treatment in HIV positive patients? |  |  |  | Do you know about HBV complications in patients with HIV? |  |  |  |
| --- | --- | --- | --- | --- | --- | --- | --- | --- | --- | --- | --- | --- |
|  | Yes<br>N (%) | No<br>N (%) | Total<br>N (%) | P | Yes<br>N (%) | No<br>N (%) | Total<br>N (%) | P | Yes<br>N (%) | No<br>N (%) | Total<br>N (%) | P |
| **Gender** |  |  |  |  |  |  |  |  |  |  |  |  |
| Female | 39 (78) | 1(50) | 40 (77) | **0.412** | 10 (83) | 30 (75) | 40 (77) | 0.433 | 9 (75) | 31 (78) | 40 (77) | **0.567** |
| Male | 11(22) | 1(50) | 12 (23) |  | 2 (17) | 10 (25) | 12 (23) |  | 3 (25) | 9 (22) | 12 (23) |  |
| **Profession** |  |  |  |  |  |  |  |  |  |  |  |  |
| Doctor | 8 (16) | 0 (0) | 8 (15) | **0.197** | 5 (42) | 3 (12) | 8 (22) | **0.009** | 6 (50) | 2 (5) | 8 (15) | **0.001** |
| Medical Technician | 9 (18) | 0 (0) | 9 (17) |  | 5 (42) | 4 (17) | 9 (25) |  | 1 (8,3) | 8 (20) | 9 (17) |  |
| Nurse | 19 (38) | 0 (0) | 19 (37) |  | 2 (16) | 17 (71) | 19 (53) |  | 4 (33,3) | 15(37,5) | 19 (37) |  |
| Counselor * |  |  |  |  |  |  |  |  | 1 (8,3) | 15(37,5) | 16 (31) |  |
| **Professional Experience** |  |  |  |  |  |  |  |  |  |  |  |  |
| 6 M-5 years | 21 (42) | 2(100) | 23 (44) | **0.269** | 4 (33) | 19 (48) | 23 (44) | **0.345** | 4 (33,3) | 19 (47) | 23 (44) | **0.568** |
| 6 a 10 years | 17 (34) | 0 (0) | 17 (33) |  | 6 (50) | 11 (28) | 17 (33) |  | 4 (33,3) | 13 (32) | 17 (33) |  |
| + 10 years | 12 (24) | 0 (0) | 12 (23) |  | 2 (17) | 10 (24) | 12 (23) |  | 4 (33,3) | 8 (20) | 12 (23) |  |

### Practices of health professionals in relation to the handling of co-infected patients

Only 15 (29%) of the participants reported screening for HBV in patients with HIV; 6 (11%) of the nurses, 4 (8%) of the medical doctors, 4 (8%) of the medical technicians and 1 (2%) of the counsellors. Only 11 (21%) reported having counselled on prevention of transmission of HBV; 4 (8%) medical doctors, 4 (8%) nurses, and 3 (5%) medical technicians. No Counsellor reported counselling to prevent the transmission of HBV infection in HIV positive patients. Only 9 (17%) provide counselling on risk behaviors (alcohol consumption, use of traditional medicine, use of intravenous drugs, use of condoms and sharing of syringes); 4 (8%) medical doctors, 3 (5%) medical technicians and 2 (4%) nurses. None of the counsellors reported counselling about risky behaviors. About what signs to look for in the patient with co-infection, the majority of participants 25 (48%) referred to jaundice. Only 4 (8%) participants mentioned having treated patients with hepatitis B “Table 5”.

**Table 5.**
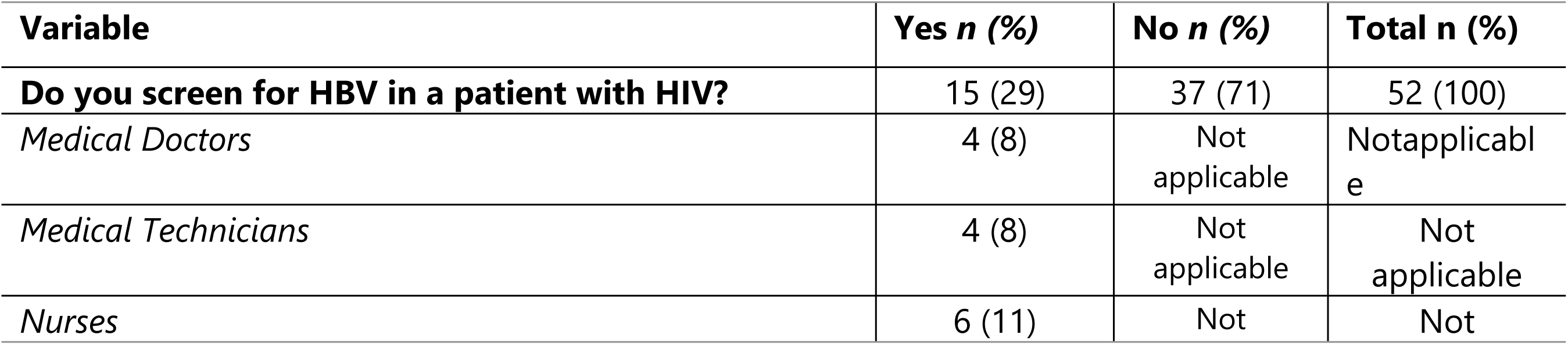
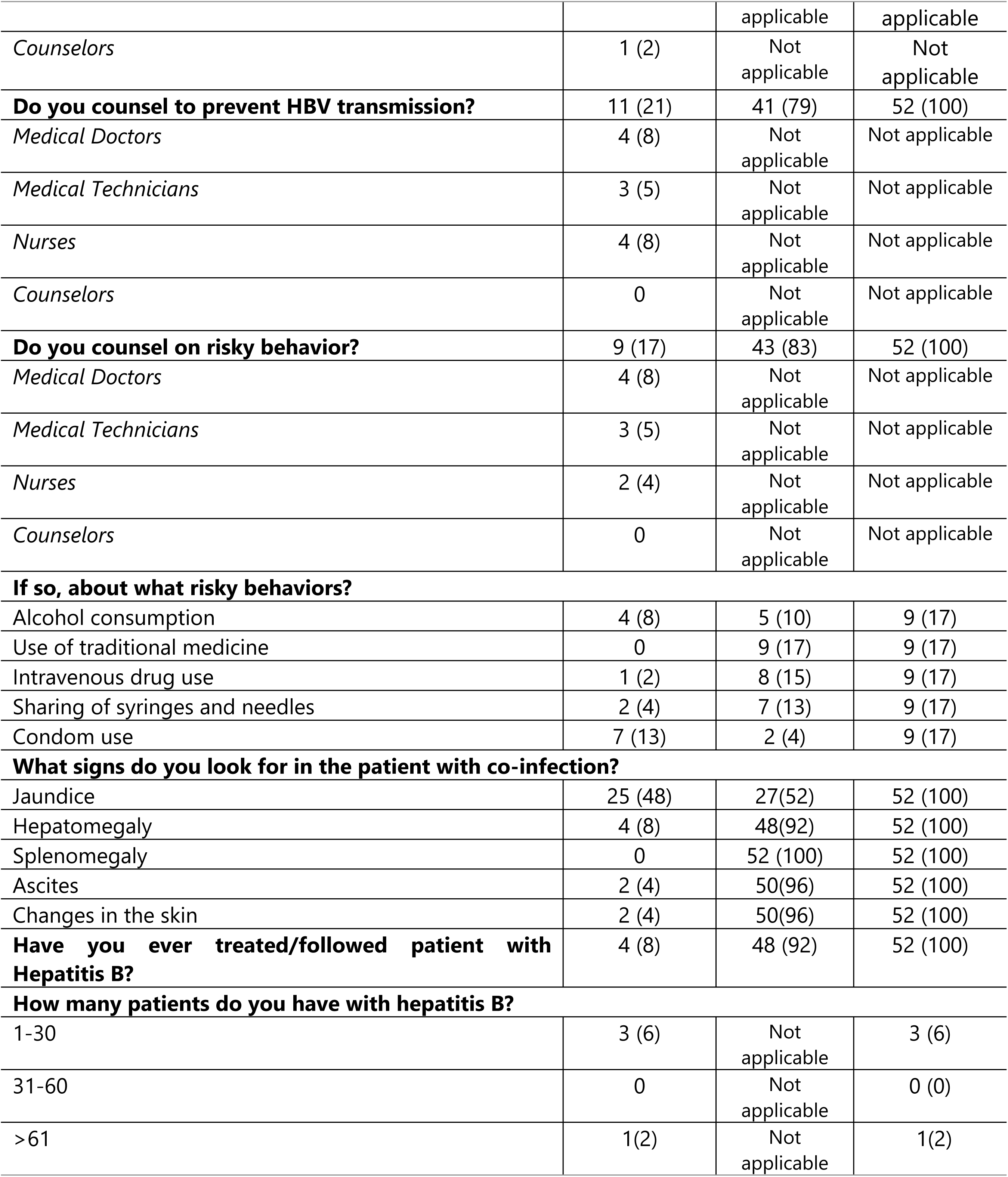
Practices of health professionals regarding HBV and HIV co-infection.

### Health professionals’ practices and sociodemographic characteristics

No statistically significant relationship was found between screening HIV patients for HBV and gender (p=0.288), professional category (0.076) and professional experience (0.056). We found an association between counselling for prevention and professional categories (p=0.028); 4 (36%) medical doctors and 4 (36%) nurses reported giving advice. It was also found that there was a statistically significant relationship between profession and counselling on risk factors (p=0.009), where 4 (44%) were medical doctors, 3 (33%) medical technicians, and 2 (22%) nurses “Table 6”.

**Table 6.**
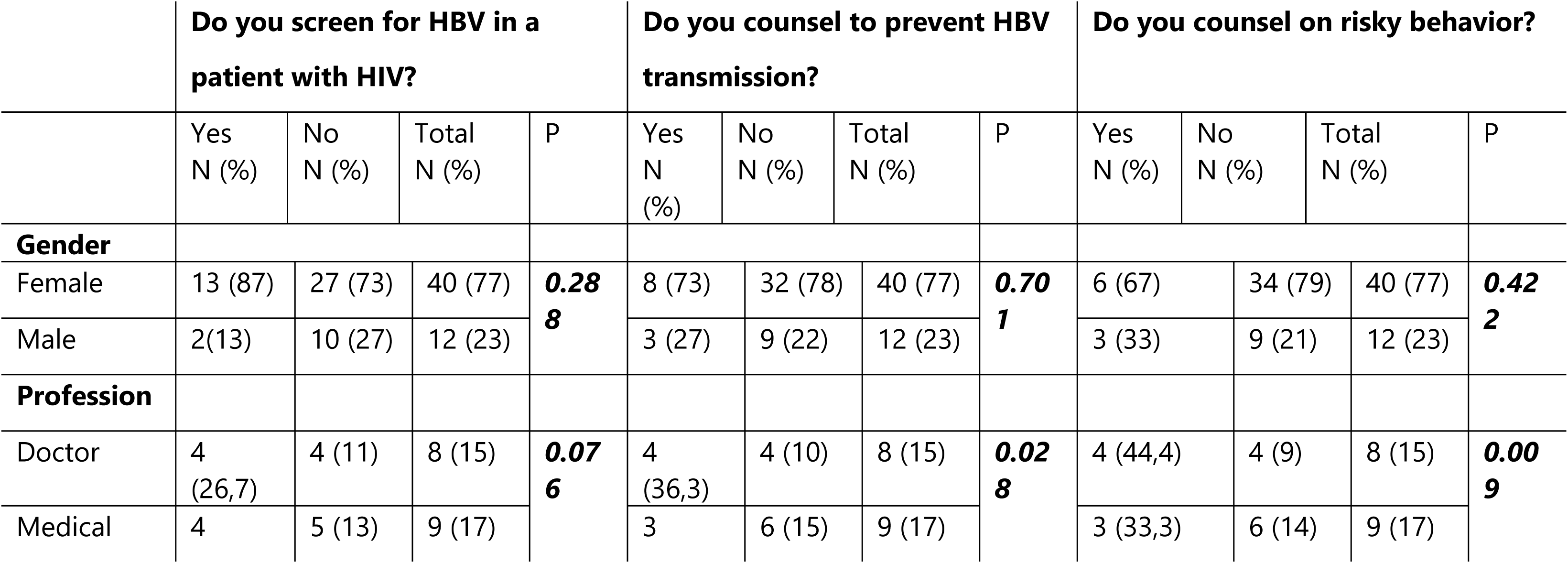
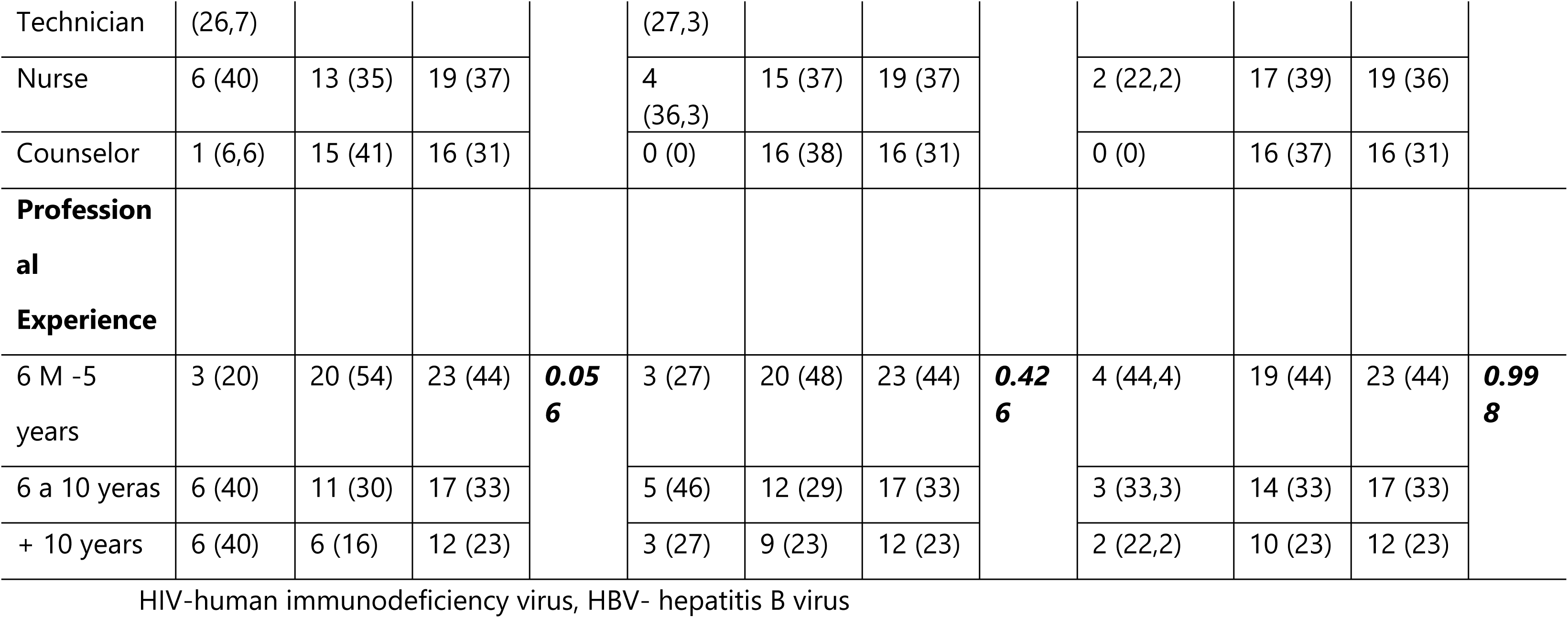
Practices and sociodemographic characteristics.

## Discussion

This study was the first carried out in Mozambique on policy review and assessment of the knowledge and practices of health professionals on HIV/HBV co-infection. We did not find studies on health professionals’ knowledge and practices concerning HIV/HBV co-infection. Additionally, we did not find policy documents specific for HIV/HBV co-infection; some guidelines was found in other policies. Most recommendations were related to the prevention of infections. Prevention measures and screening strategies for specific groups were not found, including health professionals, HIV positive patients, or other risk groups (men who have sex with men, sex workers, and intravenous drug users). The current prevention strategy is to vaccinate children under one year of age (under the Expanded Program on Vaccination) without including neonates for the prevention of HBV transmission from mother to child. Screening could help identify those who need treatment and those who would benefit from vaccination. A study in the United States has shown that strategies to screen and treat or vaccinate are cost-effective in reducing the burden of hepatitis B virus among high-prevalence populations (28).

Diagnosed HIV/HBV co-infected patients would benefit from specific guidelines on monitoring for early detection of complications and referral to a high-level facility with a specialist. The policy should be accompanied by health professionals’ awareness, training and engagement to adhere to the guidelines. Studies have described low compliance with hepatocellular carcinoma screening in HIV/HBV co-infected patients, resulting in late management (21)(22). Only one policy explicitly referred to HIV/HBV co-infection. This was a policy about the use of drugs active against HBV and HIV, a recommendation aligned with the international guidelines (29).

These results are similar to those found in a study carried out in Ethiopia (30), which revealed a lack of treatment policies and strategies for testing, diagnosis and treatment of hepatitis B. A study in Ghana (31) showed that despite the high prevalence of co-infection (12.3%), there were no policies regarding prevention, testing for HBV, or patient management in newly diagnosed HIV-positive patients. There is a need to formulate policies and identify at-risk groups who can benefit from testing, care, and treatment.

The assessment of knowledge and practices involved different categories of health professionals. We included medical doctors, medical technicians, nurses, and counsellors, as they play a critical role in the HIV/HBV co-infected care chain. Although they can intervene in any step, counsellors are more involved with prevention, nurses with screening and medical doctors and technicians with diagnosis, treatment and follow-up. Most health professionals were aware of the coexistence of HIV and HBV co-infection. When asked about transmission, preventions, diagnosis, treatment, and cure, the knowledge was insufficient. Except for a few parameters, there were no differences among all categories of health professionals participating in the study. Most professionals failed to indicate vaccinations as a preventive measure. These findings are similar to studies in India and Malawi, where health professionals had poor knowledge towards HIV/AIDS, HBV and HCV and the need for education on preventing HBV was recommended (32). It is essential that health professionals know about transmission and prevention measures for HIV/HBV co-infection, not only to counselling their patients but also to protect themselves from the risk of being infected while caring for a patient.

Diagnosing HBV infection through testing for the Hepatitis B Surface Antigen (HBsAg) was also an area of insufficient knowledge. However, there were differences between medical doctors and medical technicians who stood out compared to nurses and counselors. The better performance of medical doctors and medical technicians is expected due to their training in the diagnosis. This result is similar to that reported by Ahmad et al. (33), who showed that knowledge was associated with the professional category. Studies have described HBsAg detection as an accurate test to use in primary care not only for the diagnosis but also for patient monitoring (34)(35).

The study also showed insufficient knowledge about treatment for hepatitis B in HIV co-infected patient. Although they were better than the others, the two categories responsible for treating the patient did not perform well (7 out of 17 medical doctors and technicians did not respond correctly). Few health professionals knew the complications of HBV infection in patients with HIV. A study in Ghana to assess the prevalence of HBV in HIV patients (36) showed that health professionals had insufficient knowledge regarding handling patients with HIV/HBV co-infection, indicating the need for regular training of health professionals about hepatitis B. This lack of knowledge of health professionals can lead to the loss of opportunities for preventing, diagnosing and treating hepatitis B in the co-infected patient.

WHO (20) recommends testing for HBV in all newly diagnosed HIV-positive patients, Hepatitis B vaccination for those who are seronegative for HBV, appropriate treatment of HIV/HBV co-infected and follow-up with control of the degree of liver damage for diagnosis of cirrhosis or hepatocellular carcinoma.

Less than a third of the health professionals reported having provided counselling to prevent the transmission of infection and risk reduction. Counsellors are expected to do so, given their HIV patient care chain role. The lack of training on HIV/HBV co-infection can contribute to these malpractices. Health professionals are also not routinely screening HIV patients for HBV infection. The absence of clear guidelines and training may be a factor that impedes these activities. The cascade of care for HIV positive patients in Mozambique is well defined, with high adherence from the health professionals indicating that clear guidelines embedded in HIV care would help to improve co-infected patient care.

In the study, about 71% of health professionals had received at least one dose of the HBV vaccine. Still, only 17% know how many doses are needed to protect against infection. It shows a lack of knowledge among health professionals about the risk of HBV infection, HBV vaccine, and the importance of being fully vaccinated given the high risk of contracting the infection. We did not find a policy that recommends health professionals to be vaccinated except in case of accidental exposure. The absence of guidelines can also play a role in health professional’s vaccine uptake. WHO recommends HBV vaccination to all healthcare professionals to prevent HBV transmission in the health care unit. These results do not differ from those found in other countries in the sub-Saharan region, such as Ethiopia, Nigeria (37) and Saudi Arabia (38). Despite health professionals being aware of the vaccine, very few are vaccinated, with low vaccination rates for Hepatitis B in health professionals with even lower rates in the administration of the three doses of the vaccine.

Taking into account the WHO recommended framework of "Monitoring and Evaluation of Viral Hepatitis B and C" for the elimination of viral hepatitis as a public health threat by 2030 (14)(39); this study focused on the issue related to facilitating factors that consisted of analyzing existing policies and norms for the handling of patients co-infected with HIV/HBV and evaluating an important factor that can affect the cascade of cure, which is the knowledge and practices of health professionals in relation to handling HIV/HBV co-infection. It was observed that there are gaps in the definition of specific policies for co-infected patients and in monitoring the cascade for cure.

### Study limitation

The Health Centers included in the study are not representative of the whole country. The number of participants in the study is small and the fact that it included counsellors may have led to the observed limitation in knowledge. An analysis stratified by group was carried out to minimize this limitation. The results relating to the analysis of knowledge according to socio-demographic characteristics should be interpreted with caution given that the number of participants was low.

## Conclusions

The study did not find specific policies for preventing, diagnosing, and managing patients with HIV/HBV co-infection. There was little knowledge about HIV/HBV co-infection among health professionals, and few knew that HBV vaccination was an effective way to prevent infection. Notably, very few health professionals were vaccinated against HBV. Defining adequate policies, training the health professionals and monitoring their activity may help increase awareness, counselling for disease prevention, early diagnosis and proper treatment of HIV/HBV co-infected patients, as we observed with HIV care.

## Data Availability

All data produced in the present study are available upon reasonable request to the authors

## Acknowledgments

We would like to give special thanks to all the participants who accepted to be part of this study. All health professionals at the health facilities where the study took place for their support and collaboration during data collection.

To the local Health Centers of Mavalane, Alto-Maé, 1° de Maio and Polana Caniço, where the data were collected. To the General Hospitals of Mavalane, Polana Caniço and Chamanculo. To Mr. Hélio Martins and Mrs. Luiza Huo for helping with the statistical analysis of the data for this project.

## Author Contributions

VM and ES designed the study and wrote the first draft of the study protocol. VM, ES, LC, CN review and approved the protocol. VM collected de data. VM, ES, LC, CN supported with data quality and analysis. VM wrote the first draft of the manuscript. VM, ES, LC, CN review and approved the final version of the manuscript.

